# Refining Muscle Morphometry Through Machine Learning and Spatial Analysis

**DOI:** 10.1101/2024.12.16.24319100

**Authors:** Daisuke Ono, Honami Kawai, Hiroya Kuwahara, Takanori Yokota

## Abstract

**Aims:** Muscle morphology provides important information in differentiating the disease aetiology, but its measurement remains challenging due to the lack of an efficient and objective method. This study aimed to quantitatively refine the morphological features of muscle fibres in neuromuscular diseases using machine learning.

**Methods:** This retrospective study analysed muscle biopsy specimens on hematoxylin and eosin-staining. A machine learning-based software was developed to segment muscle fibre contours and perform automated muscle morphometry and subsequent graph theory-based spatial analysis of atrophied fibre grouping. A decision tree-based framework, LightGBM, was trained to predict underlying aetiologies based on the morphometric and spatial variables.

**Results:** The study included 100 muscle samples, including 20 normal muscles, 49 myopathies, and 19 neuropathies. The fine-tuned segmentation model, YOLOv8, achieved a mask average precision of 0.819. The muscle morphometry revealed the significance of fibre circularity. The mean circularity was higher in the myopathy group, and the SD of circularity was elevated in the neuropathy group. Although most cases were consistent with textbook findings, atypical presentations, such as dermatomyositis with angular atrophy and amyotrophic lateral sclerosis with round atrophy, were objectively documented. Spatial analysis quantified grouped atrophy, showing the potential to feature specific atrophy patterns. The LightGBM model successfully predicted the final clinical diagnosis of the myopathies and neuropathies with an accuracy of 0.852, which exceeded that of 0.808 by human annotation.

**Conclusion:** Automated muscle morphometry and spatial analysis provided the quantification of muscle morphology and atrophy patterns, which will facilitate objective and efficient investigation of neuromuscular diseases.

**Key Points:**

- This study aimed to quantitatively refine the morphological features of muscle fibres in neuromuscular diseases using machine learning-based software.
- Automated muscle morphometry successfully characterised myopathies and neuropathies, presenting the utility of circularity for their differentiation.
- Spatial analysis based on graph theory proposed quantitative definitions for grouped atrophy and perifascicular atrophy, which showed the potential to feature the grouping patterns of atrophic fibres.
- This open-source tool will facilitate objective and efficient investigation of neuromuscular diseases.

## INTRODUCTION

Muscle biopsy is performed to diagnose neuromuscular diseases. Morphological changes in muscle fibres reflect the underlying aetiology ^1–4^. In myopathies, muscle fibres become rounded, and small and large fibres are seen with significant size variation within a sample ^2,5^. In contrast, clusters of angularly atrophied fibres are observed in the muscles with neurogenic changes ^2,4,6^. Constantinides *et al* reported that the diagnostic accuracy of muscle biopsy in differentiating these two aetiologies was higher than that of electromyography (EMG) (80.9% vs. 70.7%), with 29.3% of patients showing disagreement between muscle biopsy and EMG ^7^. These findings suggest that, although the aetiological diagnosis is primarily determined by minimally invasive examinations, such as electromyography (EMG) and serum creatine kinase (CK), muscle morphology provides an additional diagnostic value with certain patients, for instance, those with a differential diagnosis of inclusion body myositis (IBM) and motor neuron disease ^8^.

To objectively characterise morphological changes, early studies manually measured muscle fibre diameters and reported important basic findings, such as changes in fibre size related to sex and age in normal muscles ^9,10^. Subsequent studies further detailed the morphological changes in specific diseases with a relatively small number of cases ^5,6,11–13^. However, conducting more comprehensive studies with larger datasets has been hindered by the extremely time-consuming and laborious nature of manual morphometry. As a result, the current aetiological diagnosis is largely based on subjective evaluation, and an efficient and objective method for qualitatively measuring muscle fibre morphology is warranted.

Automated approaches for muscle morphometry have been explored, from rule-based contour detection tools ^14–23^, to machine learning-based approaches ^24–29^. Those models successfully segmented the contours of muscle fibres using hematoxylin and eosin staining (H&E), immunofluorescence, or ATPase activity ^30^. However, most of these models specifically targeted animal muscles in experimental settings or normal human muscles. Although several studies have performed automated morphometry on human muscles with muscular dystrophy ^16,22,23^, no study has yet focused on characterising myopathic and neurogenic changes with automated muscle morphology.

In this study, we aimed to quantitatively refine the morphological features of muscle histology in neuromuscular diseases. To achieve this, we developed a machine-learning-based software for automated muscle morphometry. This open-source software inputs a whole slide image (WSI) of a biopsied muscle, automatically performs morphometric and spatial analyses, and analyses the underlying aetiology of the neuromuscular disease.

## MATERIALS AND METHODS

### Patient enrollment and clinicopathologic evaluation

This was a retrospective, clinicopathological study enrolling men and women aged 18 years or older who underwent muscle biopsy at the Institute of Science Tokyo Hospital between January 1, 2015, and April 30, 2022. The medical records and pathology reports were retrospectively reviewed. To avoid circular logic in the definition of myopathic and neurogenic changes, the aetiological categories of myopathy and neuropathy were based on the final clinical diagnoses rather than the pathological diagnoses. Among the samples with morphological changes on H&E-stained slides, those with the final clinical diagnosis of myopathy or peripheral neuropathy were assigned to the myopathy or neuropathy group, respectively. Amyotrophic lateral sclerosis (ALS) with lower motor neuron signs was included in the neuropathy group in this study. Samples without morphological changes were classified as the normal group. Specimens with severe artefacts, those with few or no muscle fibres present on the slide (end-stage muscle), and those with comorbid myopathy and neuropathy were included to train a segmentation model to strengthen the model performance, but were thereafter excluded from clinicopathologic analysis.

The clinical diagnoses of ALS and autoimmune diseases were based on the standard diagnostic criteria ^31–34^, while the diagnosis of idiopathic inflammatory myopathies (IIMs) was based on diagnostic criteria at the time of evaluation. Some patients who presented with no myositis-related antibody positivity or no characteristic findings, such as IBM, immune-mediated necrotizing myopathy (IMNM), dermatomyositis (DM), or antisynthetase syndrome (ASS), were classified as having polymyositis (PM), which is currently not considered an independent disease ^35^.

### Sample preparation and scanning slides

Biopsied samples were immediately frozen at -80 °C after the procedure, then sectioned at 10 μm, stained with Hematoxylin (#131-09665; FUJIFILM Wako Pure Chemical Corporation, Osaka, Japan) and 1%Eosin Y (#058-00062; FUJIFILM Wako Pure Chemical Corporation, Osaka, Japan). All slides were scanned at 20 x magnification, and digitised to WSIs with 0.2730 µm / pixel resolution with Olympus VS200 (Olympus Life Science, Tokyo, Japan).

### Machine Learning for muscle fibre segmentation

Square tiles of 2000 × 2000 pixels were randomly sampled from WSIs, specifically five tiles from each of the 100 cases. After excluding 33 tiles that contained only background without muscle tissue, 467 tiles were obtained. Each contour of the muscle fibres on the tiles was annotated by two neurologists (DO and HK) using Label Studio (https://github.com/HumanSignal/label-studio). The dataset was divided into the training, validation, and test datasets at a ratio of 0.70:0.15:0.15. An instance segmentation model, yolov8l-seg from YOLOv8.0.91, was fine-tuned on the training and validation datasets using Google Colab Pro (GPU: NVIDIA A100-SXM4-40GB, Python-3.10.11, torch-2.0.0+cu118). The model performance was evaluated on the holdout test dataset using box average precision (AP, IoU=0.50:0.95) for muscle fibre detection and mask AP (IoU=0.50:0.95) for segmentation. These indices are scored from 0 to 1, with values closer to 1 indicating a better performance. For morphometry, 2000 pixel square tiles with 300 pixel overlapping margins were generated and processed, as previously reported ^36^. After the segmentation of each border by the fine-tuned model, overlapping objects were identified and filtered based on the thresholds of their areas and intersection areas.

### Morphometry

The basic properties of fibres, such as cross-sectional area (CSA), circularity, and eccentricity, were measured as previously reported^36^. Since obliquely-sectioned muscle fibres disturb the morphometric analysis, we excluded muscle fibres with eccentricity greater than 5. Muscle fibres were classified based on the means and standard deviations (SD) calculated from the minimal change group, as follows: large fibres (log(area) > mean + SD), small fibres (log(area) < mean + SD), round fibres (circularity > mean + SD), and angular fibres (circularity < mean - SD).

### Spatial analysis

Graph theory was applied to quantify the grouping of atrophic fibres. Each center of a small fibre is represented as a node. Edges were drawn between nodes within 50 µm of each other. The distance was determined with reference to previous reports stating that the mean average diameter of normal adult muscle fibres was 40-60 µm ^3,4,9^. We subsequently applied the community Louvain algorithm from the Python library NetworkX 2.8.8 to detect clusters consisting of at least three fibres. The ratio of the number of small fibres in the clusters to the total number of grouped fibres (%), mean number of nodes per cluster, and edges per node were computed.

### Prediction model

To develop an automated prediction model for the differentiation of myopathy and neuropathy, we applied the Light Gradient Boosting Machine (LightGBM), a decision-tree-based ensemble learning framework ^37^. The model was trained using the morphological and spatial parameters obtained from both groups. Using 10-fold cross-validation, the performance on the test dataset was assessed by computing the accuracy, macro-averaged precision, and area under the receiver operating characteristic curve (AUC). The key parameters employed in the LightGBM model configuration included: ‘num_classes’: 3, ‘metric’: ‘multi_logloss, ‘num_leaves’: 31, ‘feature_fraction’: 0.9, ‘learning_rate’: 0.05, ‘bagging_fraction’: 0.8, ‘bagging_freq’: 5, ‘verbose’: -1, ‘max_depth’: 10, and ‘n_estimators’: 10000. To compare predictions by the LightGBM model with human annotations, H&E slides were reviewed and classified as myopathic or neurogenic changes by a board-certified neurologist, DO, blinded to clinicopathologic information. Specimens with round fibres and their size variations were considered myopathic changes, whereas those with angular fibres and their clusters were considered neurogenic changes^1–4^.

### Software development and code availability

We implemented a machine learning model for muscle morphometry into software that inputs the WSI of a biopsied muscle on H&E staining, and outputs morphometric and spatial analysis. This model has been packaged as an open-source Python library, and is available at https://github.com/onnonuro/mmmetry.git with a Jupyter Notebook tutorial.

### Statistical analysis

All statistical analyses were performed using Python and its libraries scikit-learn and scipy. Statistical differences between pathological categories were evaluated using the chi-square test for categorical variables, and the Kruskal-Wallis test for continuous variables, followed by the pairwise Steel-Dwass test as a post-hoc analysis for those with significant differences. Statistical significance was set at P < 0.05. Data are presented as the median (interquartile range), unless otherwise noted.

## RESULTS

### Patients characteristics and segmentation model

During 2015-2022, 100 patients underwent muscle biopsy, including 20 normal muscles, 49 myopathies, and 19 neuropathies (Table 1). The diagnoses included 34 cases of IIMs, 7 cases of muscular dystrophy, 9 cases of vasculitic neuropathy, and 4 cases of ALS. We fine-tuned the instance segmentation model YOLOv8 using 467 tiles randomly sampled from 100 digitised slides (Figure 1). The model performance was 0.819 for the mask AP for fibre segmentation, and 0.813 for the box AP for fibre detection.

**FIGURE 1.**
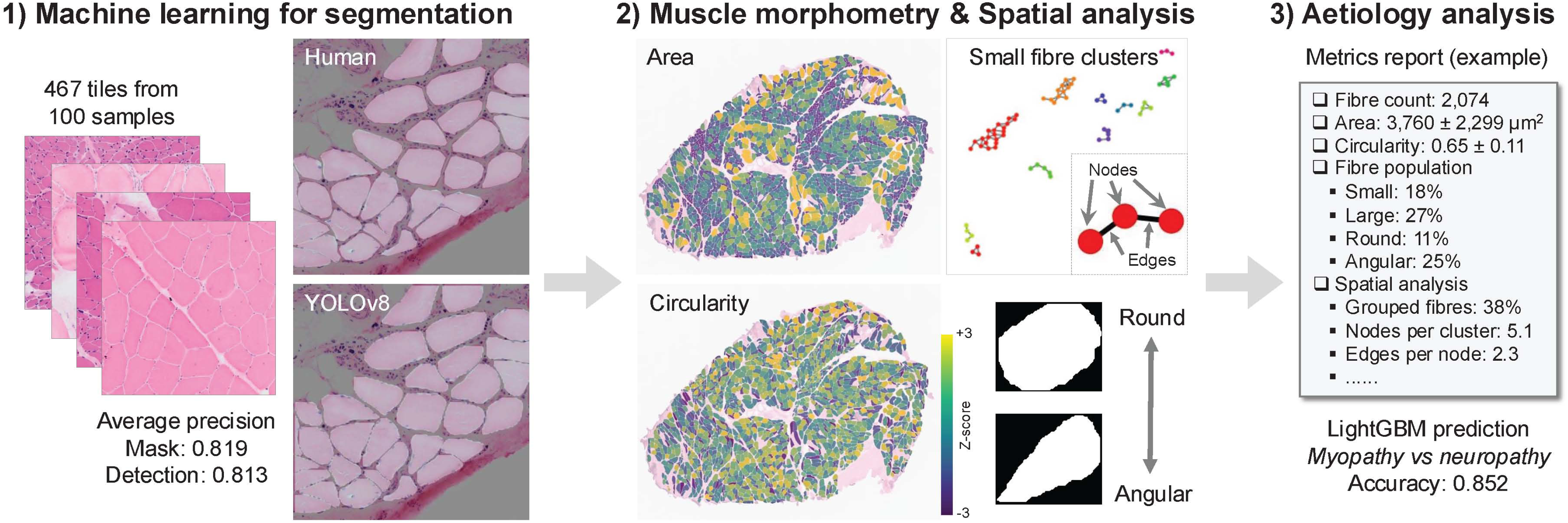
Workflow of the study. For automated segmentation of muscle fibres, a machine learning model, YOLOv8, was fine-tuned with 467 tiles from 100 muscle biopsy specimens, achieving a mask average precision of 0.819. 2) The fine-tuned model quantified morphological features, such as cross-sectional area and circularity. Small fibre clusters were detected based on graph theory. 3) Metrics from morphometry and spatial analysis were used to train a LightGBM model, achieving an accuracy of 0.852 in distinguishing myopathies from neuropathies. LightGBM, light gradient boosting machine; YOLOv8, you only look once version 8.

**Table 1.**
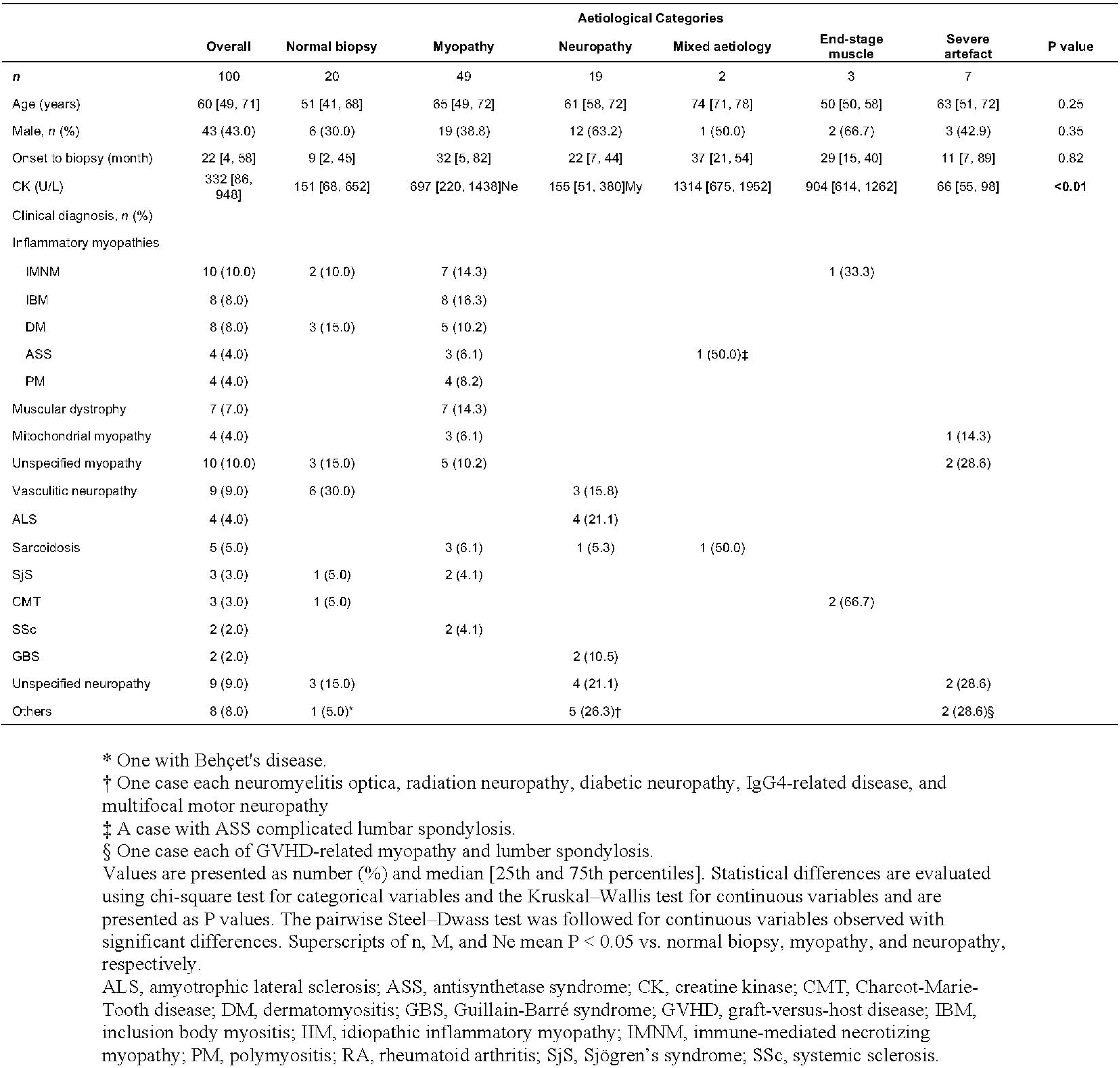
Patient characteristics.

|  | Aetiological Categories |  |  |  |  |  |  | P value |
| --- | --- | --- | --- | --- | --- | --- | --- | --- |
|  | Overall | Normal biopsy | Myopathy | Neuropathy | Mixed aetiology | End-stage muscle | Severe artefact |  |
| *n* | 100 | 20 | 49 | 19 | 2 | 3 | 7 |  |
| Age (years) | 60 [49, 71] | 51 [41, 68] | 65 [49, 72] | 61 [58, 72] | 74 [71, 78] | 50 [50, 58] | 63 [51, 72] | 0.25 |
| Male, *n* (%) | 43 (43.0) | 6 (30.0) | 19 (38.8) | 12 (63.2) | 1 (50.0) | 2 (66.7) | 3 (42.9) | 0.35 |
| Onset to biopsy (month) | 22 [4, 58] | 9 [2, 45] | 32 [5, 82] | 22 [7, 44] | 37 [21, 54] | 29 [15, 40] | 11 [7, 89] | 0.82 |
| CK (U/L) | 332 [86, 948] | 151 [68, 652] | 697 [220, 1438]<sup>Ne</sup> | 155 [51, 380]<sup>My</sup> | 1314 [675, 1952] | 904 [614, 1262] | 66 [55, 98] | **<0.01** |
| Clinical diagnosis, *n* (%) |  |  |  |  |  |  |  |  |
| Inflammatory myopathies |  |  |  |  |  |  |  |  |
| IMNM | 10 (10.0) | 2 (10.0) | 7 (14.3) |  |  | 1 (33.3) |  |  |
| IBM | 8 (8.0) |  | 8 (16.3) |  |  |  |  |  |
| DM | 8 (8.0) | 3 (15.0) | 5 (10.2) |  |  |  |  |  |
| ASS | 4 (4.0) |  | 3 (6.1) |  | 1 (50.0)<sup>‡</sup> |  |  |  |
| PM | 4 (4.0) |  | 4 (8.2) |  |  |  |  |  |
| Muscular dystrophy | 7 (7.0) |  | 7 (14.3) |  |  |  |  |  |
| Mitochondrial myopathy | 4 (4.0) |  | 3 (6.1) |  |  |  | 1 (14.3) |  |
| Unspecified myopathy | 10 (10.0) | 3 (15.0) | 5 (10.2) |  |  |  | 2 (28.6) |  |
| Vasculitic neuropathy | 9 (9.0) | 6 (30.0) |  | 3 (15.8) |  |  |  |  |
| ALS | 4 (4.0) |  |  | 4 (21.1) |  |  |  |  |
| Sarcoidosis | 5 (5.0) |  | 3 (6.1) | 1 (5.3) | 1 (50.0) |  |  |  |
| SjS | 3 (3.0) | 1 (5.0) | 2 (4.1) |  |  |  |  |  |
| CMT | 3 (3.0) | 1 (5.0) |  |  |  | 2 (66.7) |  |  |
| SSc | 2 (2.0) |  | 2 (4.1) |  |  |  |  |  |
| GBS | 2 (2.0) |  |  | 2 (10.5) |  |  |  |  |
| Unspecified neuropathy | 9 (9.0) | 3 (15.0) |  | 4 (21.1) |  |  | 2 (28.6) |  |
| Others | 8 (8.0) | 1 (5.0)<sup>*</sup> |  | 5 (26.3)<sup>†</sup> |  |  | 2 (28.6)<sup>§</sup> |  |

\* One with Behçet's disease.
† One case each neuromyelitis optica, radiation neuropathy, diabetic neuropathy, IgG4-related disease, and multifocal motor neuropathy
‡ A case with ASS complicated lumbar spondylosis.
§ One case each of GVHD-related myopathy and lumbar spondylosis.
Values are presented as number (%) and median [25th and 75th percentiles]. Statistical differences are evaluated using chi-square test for categorical variables and the Kruskal–Wallis test for continuous variables and are presented as P values. The pairwise Steel–Dwass test was followed for continuous variables observed with significant differences. Superscripts of n, M, and Ne mean $P < 0.05$ vs. normal biopsy, myopathy, and neuropathy, respectively.
ALS, amyotrophic lateral sclerosis; ASS, antisynthetase syndrome; CK, creatine kinase; CMT, Charcot-Marie-Tooth disease; DM, dermatomyositis; GBS, Guillain-Barré syndrome; GVHD, graft-versus-host disease; IBM, inclusion body myositis; IIM, idiopathic inflammatory myopathy; IMNM, immune-mediated necrotizing myopathy; PM, polymyositis; RA, rheumatoid arthritis; SjS, Sjögren's syndrome; SSc, systemic sclerosis.

### Muscle morphometry

In total, 88 muscle WSIs from the normal, myopathy, and neuropathy groups were subjected to automated muscle morphometry and spatial analysis (Table 2). First, we measured the CSA and calculated the circularity of each muscle fibre in the normal muscles, with the results revealing a significant variation in CSA distribution in each normal muscle (Figure 2A,B). Neither CSA nor circularity in the normal muscles showed a correlation with age or sex (Table S1). The mean and SD of the CSA did not show any difference between the aetiological categories (P = 0.74, and 0.12, respectively). In contrast, the circularity in each normal muscle showed less variability (Figure 2C,D). The mean circularity was higher in the myopathy group (P < 0.05). The SD of circularity was elevated in patients with neuropathy (P < 0.05). Cutoffs were set at 1504/4740 µm^2^ for CSA, and 0.588/0.788 for circularity, based on the mean ± SD of the normal biopsy group. The small/large and/or round/angular fibre populations were defined using these cutoffs. These findings generally corresponded to textbook findings, such as increased small round fibres in myopathy, and increased small angular fibres in neuropathy (Figure 3A-C). However, some patients do not follow this general rule. For example, a muscle sample from an anti-MDA5 antibody-positive DM patient showed type 2 fibre-dominant angular atrophy (Figure 3C,D and Figure S1), while another from an ALS patient showed round fibre-dominant atrophy (Figure 4A,B). Subanalysis of the IIM revealed that the IBM presented fewer fibres and increased the population of large round fibres (Table S2).

**FIGURE 2.**
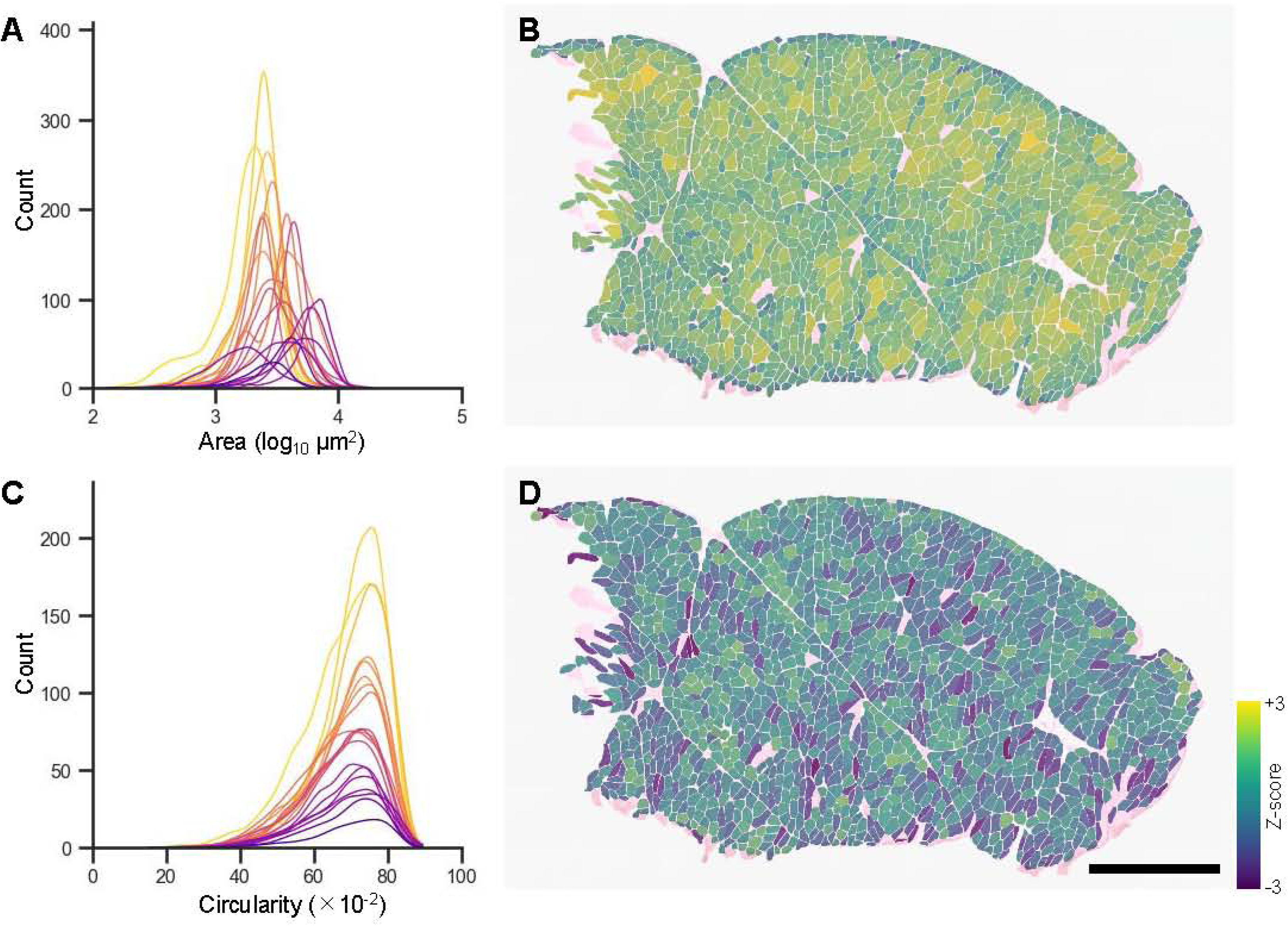
Distribution of cross-sectional area and circularity in the normal muscle. (A, C) Distribution of cross-sectional area (A) and circularity (C) in normal biopsied muscles. Each curve, coloured based on the number of muscle fibres, from deep purple (fewest) to yellow (most), represents a kernel density estimation on counts in 100 bins in individual samples. (B, D) Representative images of normal biopsied muscle on H&E staining. Each muscle fibre is coloured with the corresponding Z-score of the cross-sectional area (B) or circularity (D). Scale bars: 1 mm.

**FIGURE 3.**
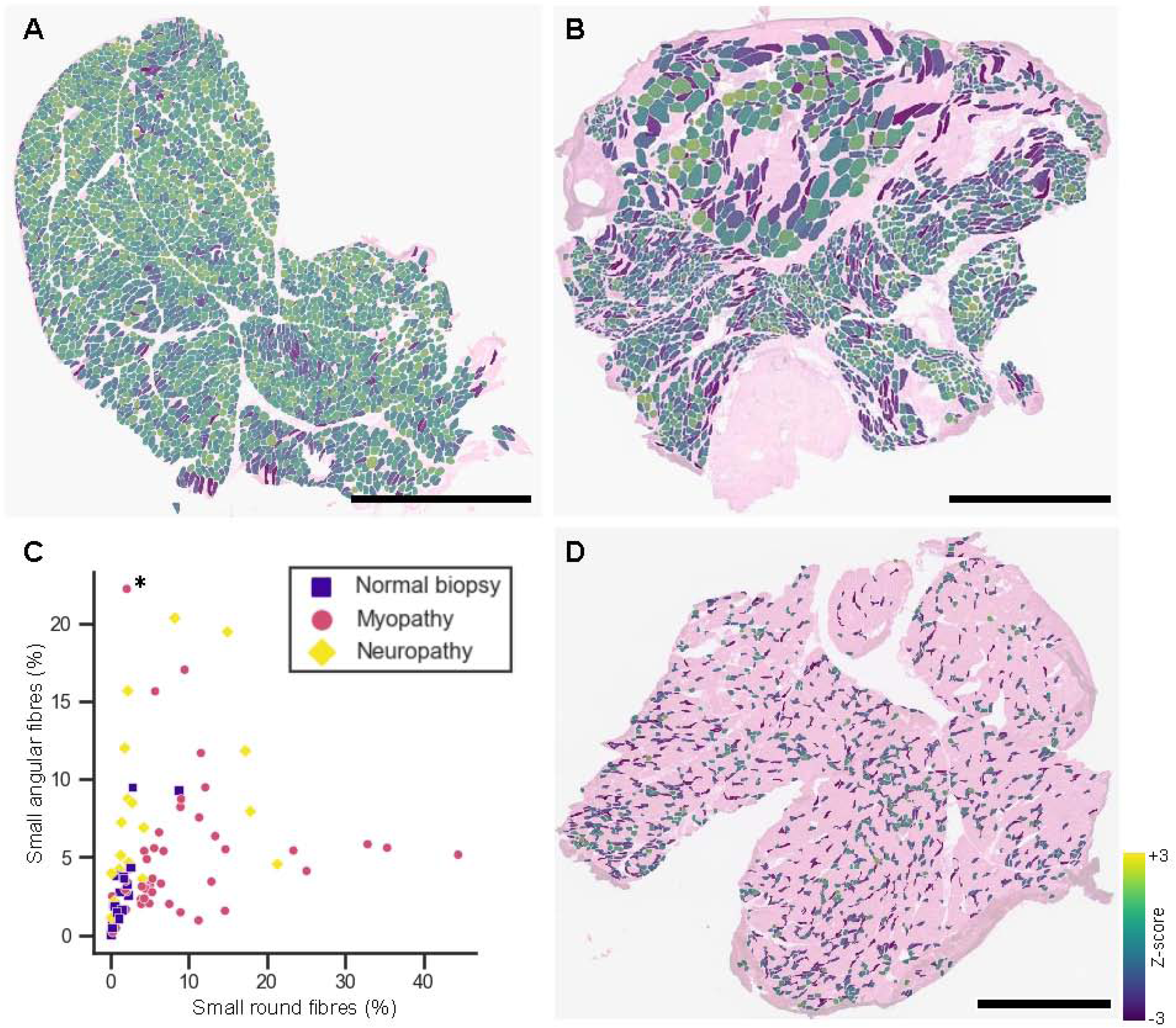
Morphometric changes in myopathy and neuropathy. (A,B) Representative H&E staining images with colour mapping of the Z-score of circularity. Small round fibres are predominantly detected in a muscle of anti-TIF1γ antibody positive dermatomyositis (A). Angular fibres are observed existing in grouped small fibres, indicating neurogenic changes due to Sjögren’s syndrome (B). (C) A scatter plot of small round and small angular fibre populations labeled with aetiological categories. Tendencies of round fibre-dominant atrophy in myopathies and angular fibre-dominant atrophy in neuropathies are observed with limited exceptions. The case shown in D is indicated by an asterisk. (D) H&E-staining of the sample from a patient with anti-MDA5 antibody positive dermatomyositis, with small fibres coloured based on circularity. Angular fibre-dominant atrophy is observed, which is atypical for myopathy. Scale bars: 2 mm in A, 1 mm in B and C. MDA5, melanoma differentiation–associated gene 5; TIF1γ, transcriptional intermediary factor 1γ.

**FIGURE 4.**
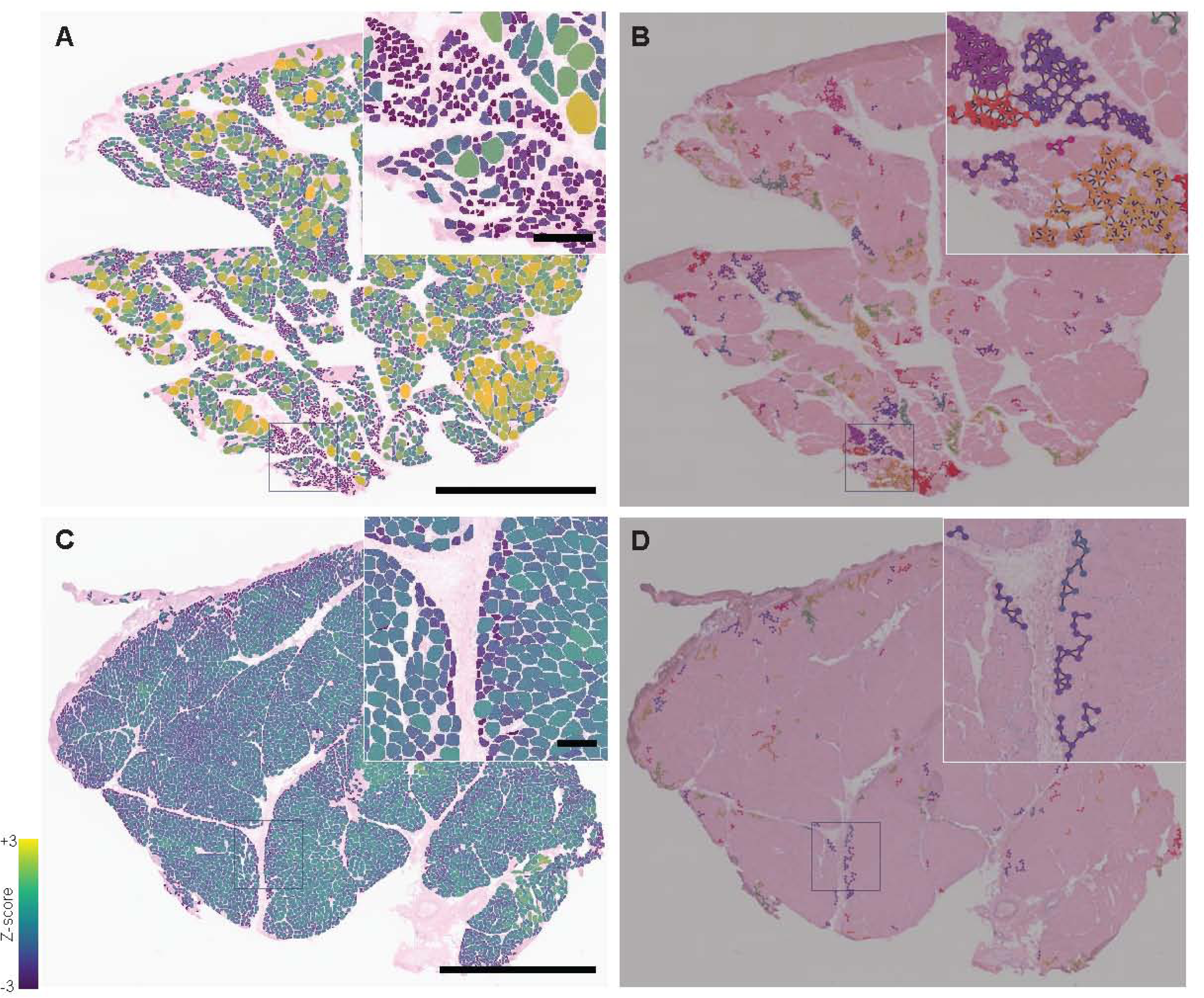
Quantification of grouped atrophy based on graph theory. Visualization of different grouping patterns observed in biopsied muscles with ALS (A, B) and anti-Jo1 antibody-positive ASS (C, D). Whole-slide images of H&E staining were mapped with the Z-score of the cross-sectional area (A, C), as well as with clusters determined by graph theory (B, D). (A, B) Large grouped atrophy is observed in ALS. Most of the atrophic fibres were round, which usually indicates myopathic changes. (C, D) In contrast, perifascicular necrosis is observed in ASS with small round fibres lining the fascicles. These grouping patterns of small fibres are quantitatively differentiated by the average number of edges per node, which is greater in ALS than in ASS cases (3.7 vs 2.3, respectively), as we as the average number of nodes per cluster (10.9 vs 6.1). Scale bars: 2 mm in A and B, 200 µm in insets in A and B. ALS, amyotrophic lateral sclerosis; ASS, antisynthetase syndrome.

**Table 2.**
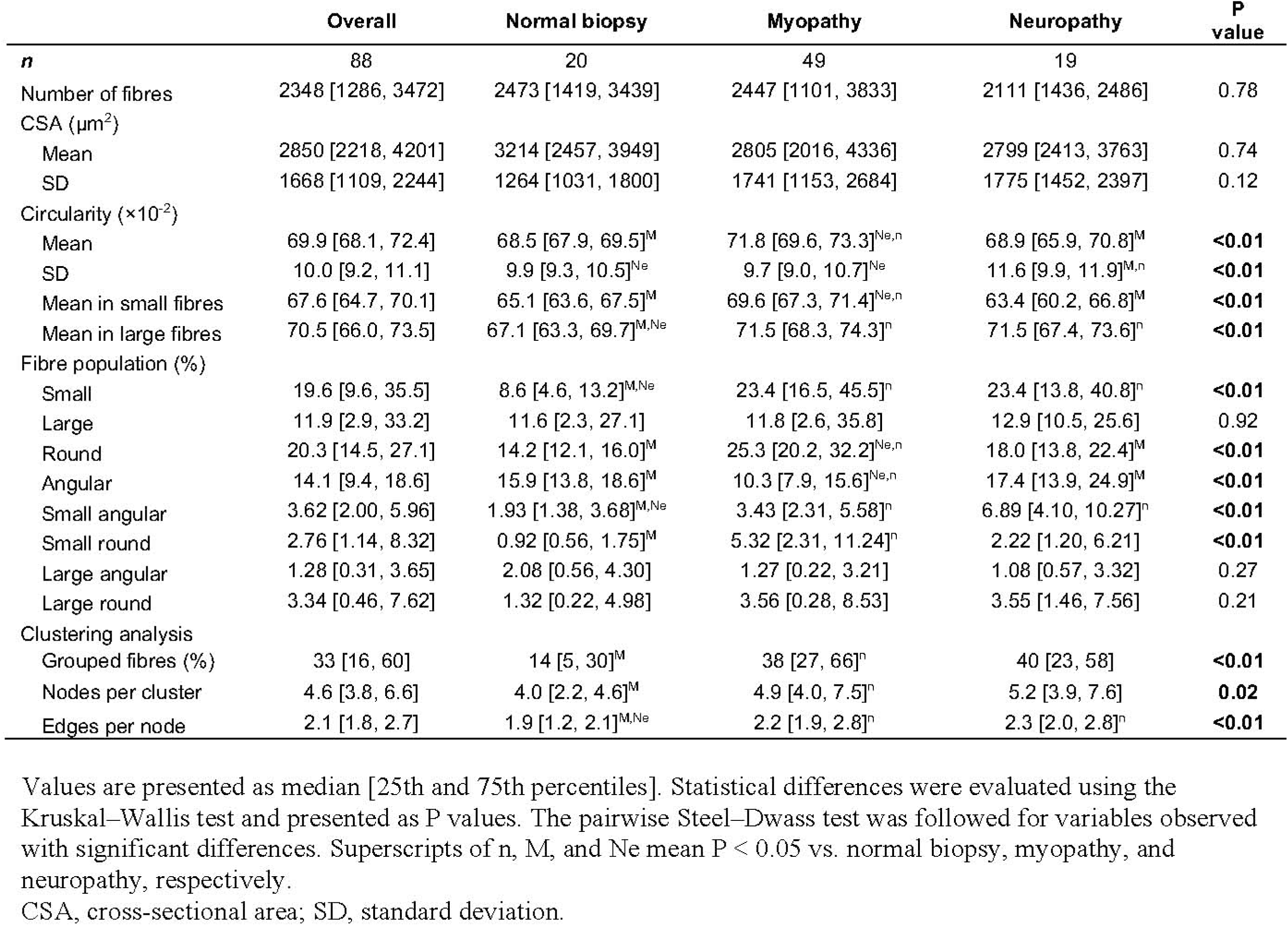
Muscle morphometry and spatial analysis.

### Spatial analysis based on graph theory

To quantify grouped atrophy using graph theory, we defined a cluster as at least 3 small fibres neighboring within 50 µm between the centers (Table 2 and Figure 4). Grouped small fibres and nodes per cluster (fibres per group) were increased in the myopathy group than in the normal group (P < 0.05). Edges per node (connections per fibre) was found to have increased in both myopathy and neuropathy groups compared to normal biopsy (P < 0.05), but no significant difference was found between the two. Despite the lack of statistical significance, these variables showed the potential utility in specific patients. For example, a muscle from a patient with ALS demonstrated small fibre atrophy, which is atypical for neurogenic disease, but the number of edges per node was high at 3.7, which was considered to reflect grouped atrophy (Figure 4A,B). Another example is perifascicular necrosis, which was observed in a muscle from a patient with ASS (Figure 4C,D) ^38^. The number of edges per node in this case was 2.3, nearly 2, which indicates that each small fibre with two neighboring fibres was lined up according to the fascicles.

### Prediction model for myopathy and neuropathy

The LightGBM was trained to predict the final clinical diagnoses of myopathy and neuropathy. First, we used only morphometric parameters, subsequently obtaining an accuracy of 0.823 ± 0.123 (mean ± SD from 10-fold cross-validation), macro-averaged precision of 0.846±0.180, and an AUC of 0.784 ± 0.187. Next, we included parameters from spatial analysis, and the model performance increased to an accuracy of 0.852 ±0.090, macro-averaged precision of 0.863±0.118, and AUC of 0.817 ±0.165, which were also higher than those by human evaluation with an accuracy of 0.808, macro-averaged precision of 0.766, and AUC of 0.738. Mislabeling by the LightGBM model included three samples of ALS with the prediction of myopathy (Figure 5A). One case of DM (Figure 3C, D) with angular fibre-dominant atrophy was misdiagnosed as neuropathy. The mean future importance for each value suggested that the mean circularity in small fibres, the population of large angular fibres, and the SD of circularity were important discriminators (Figure 5B).

**FIGURE 5.**
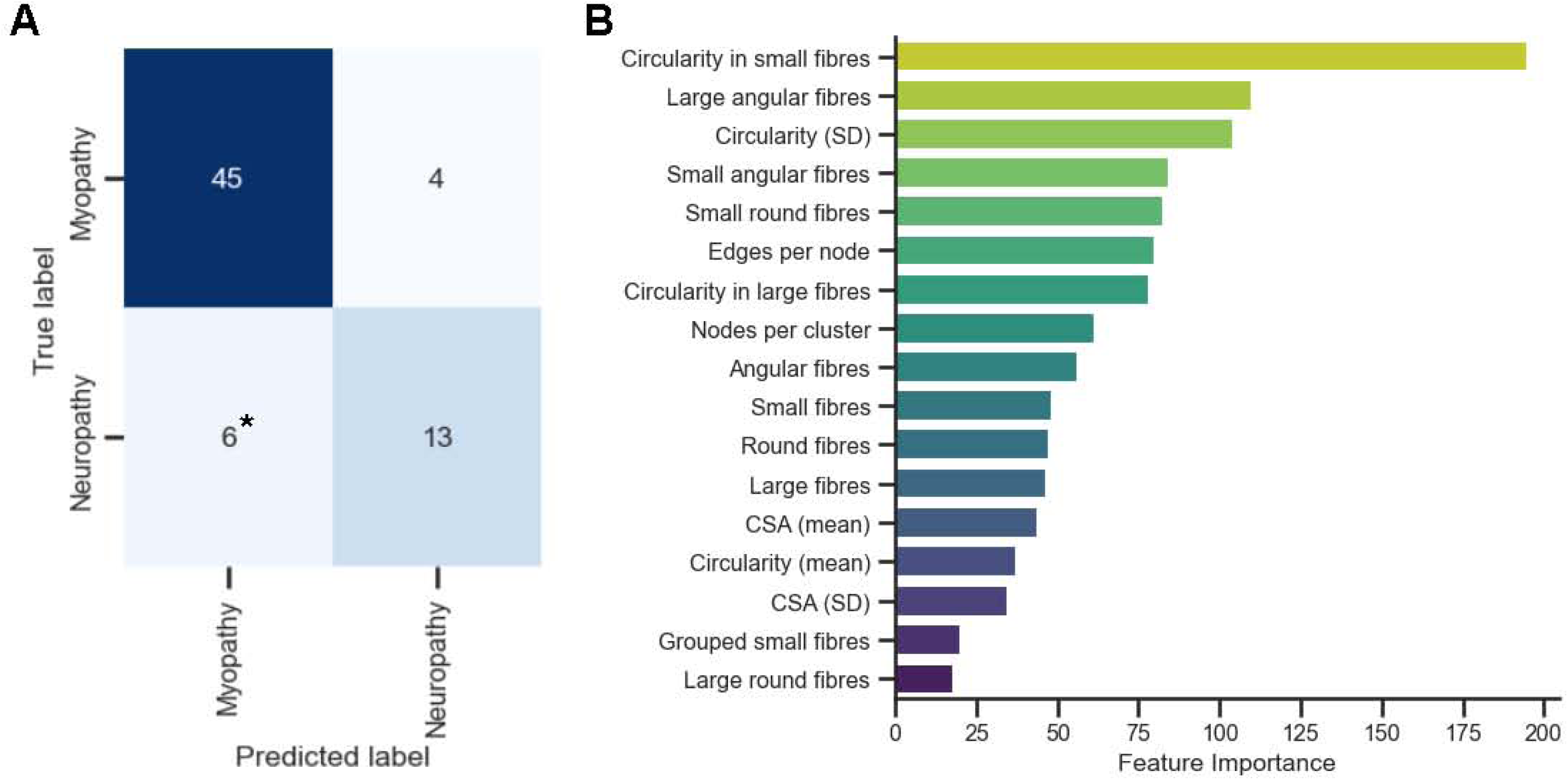
Machine learning-based prediction of aetiologies. A decision tree-based ensemble learning framework, Light Gradient Boosting Machine is applied to determine muscle aetiologies. The trained model predicts myopathy or neuropathy on a biopsied muscle based on morphological and spatial parameters with accuracy of 0.852 ±0.090 (mean ± SD), and an AUC of 0.817 ±0.165. (A) Confusion matrix obtained from the 10-fold test results. Three samples of amyotrophic lateral sclerosis are mislabeled as myopathy (*). (B) The mean future importance of each input value is visually represented. AUC, area under the curve; CSA, cross-sectional area; SD, standard deviation.

## DISCUSSION

In the current study, we developed an open-source software to perform automated muscle morphometry and spatial analysis of H&E-stained muscle WSIs. This software enabled the comprehensive investigation of human muscles with a variety of neuromuscular diseases, allowing us to propose objective parameters to characterise neurogenic and myogenic features.

We have developed a robust muscle morphometry pipeline for human samples. The segmentation model for muscle fibres was trained using 467 patches randomly sampled from 100 muscle specimens. The performance of the model on the holdout test dataset was satisfactory for our purposes, achieving a mask AP of 0.819 for fibre segmentation, and a box AP of 0.813 for fibre detection. Because we retrospectively enrolled as many serial cases as possible, the muscles used for training encompassed a wide variety of diagnoses, patient ages, disease stages, and staining conditions. Previous machine-learning-based models for muscle morphometry were trained using muscles from animal models or normal humans ^24–29^. The diversity of our training dataset is a significant strength of the current model. Furthermore, our open-source software was designed for ease of use by both clinicians and pathologists. We anticipate that the software will be applied to standardise muscle biopsy assessments across facilities for future clinical trials.

The current study demonstrates the utility of circularity in differentiating disease aetiologies. To date, muscle morphometry in human neuromuscular diseases has primarily focused on the CSA and its distribution ^5,6,9–13^. However, wide variability in CSA has been reported depending on the biopsied muscle, age, sex, and exercise level, making comprehensive research with human biopsied samples difficult ^9,10,39,40^. Our analysis showed that the mean and SD of CSA in each normal muscle varied widely, and did not differ between aetiologies (Figure 2A,B, and Table 2). In contrast, the mean and SD of circularity showed less variability throughout the normal muscles, demonstrating significant differences between the aetiologies (Figure 3C and Table 2). Several studies have measured the circularity of muscle fibres in normal human or mouse muscles ^24,40^; however, the current study is noteworthy in that it successfully demonstrated their utility for differentiating disease aetiologies in biopsied human specimens.

This study quantitatively confirmed the practicality of the textbook findings, but also cautioned regarding potential issues with over-adapting these rules. Overall, we observed an increased population of small round fibres in myopathy and that of small angular fibres in neuropathy, which is consistent with the textbook findings ^1–4^. This study also objectively identified a small number of exceptions to these general rules that were not overlooked (Figure 3C). Some patients with ALS demonstrated small round atrophy (Figure 4A,B), which has been reported, particularly those with higher serum CK levels ^41,42^. In the representative case, the nodes per cluster increased to 10.9, indicating the large group atrophy characteristic of neurogenic changes. Although no significant differences were observed between the myopathy and neuropathy groups (Table 2), this metric may still be useful for describing large group atrophy in individual cases of neuropathy.

In the present study, muscle morphometry also revealed that some cases of myopathy had more small angular fibres than small round fibres (Figure 3C,D) ^7,12^. Notably, the representative case of anti-MDA5 antibody-positive DM exhibited type 2 fibre-dominant angular atrophy, consistent with previous reports on other myopathies such as GNE myopathy. ^43^ The current study included a limited number of cases with type 2 fibre-dominant angular atrophy, making detailed analysis challenging. However, the size of grouped atrophy, represented by nodes per cluster, might be useful in differentiating it from neurogenic changes in individual cases. A subanalysis of IIM revealed an increased large fibre population in IBM, with a median interval from onset to biopsy of 71 months (Table S2), suggesting that many cases were biopsied in the chronic phase. Although compensatory hypertrophy in the chronic phase of IBM may be its characteristic finding, the small number of cases precludes definitive conclusions.

This study proposes the quantification of grouped atrophy based on graph theory. We defined grouped atrophy as a cluster of at least 3 small neighboring fibres within 50 μm between centers, identified by the Community Louvain algorithm. These parameters were consistent with subjective assessments (Figure 4), but comparisons of spatial variables between the myopathy and neuropathy groups revealed no differences. The current study included samples with mild morphological changes, and then the relatively low prevalence of grouped atrophy in both groups might have masked statistical power. However, the utility of these parameters was supported by the fact that adding spatial parameters increased the prediction performance in the LightGBM model (Figure 5). Furthermore, we expect the spatial parameters would help to understand grouping patterns of atrophied fibres in specific cases. The number of nodes per cluster means the number of fibres in each cluster, which defines the size of the grouped atrophy. The edges per node represent the number of neighboring fibres in each fibre, which explains how atrophied fibres are grouped on a slide. If the value is close to 2, it means that each atrophied fibre lines up with two neighbors, which potentially corresponding to perifascicular atrophy or necrosis ^11,38^.

We trained the LightGBM to predict the final clinical diagnoses based on morphological and spatial parameters, achieving an accuracy of 0.852 ±0.090, macro-averaged precision of 0.863±0.118, and an AUC of 0.817 ±0.165, which were more accurate than human annotation. This study included imbalanced labels of neuropathy (n = 19) and myopathy (n = 49), but the high macro-averaged precision ensures robust performance across both labels by evaluating minor and major labels equally. Our model incorrectly predicted three ALS samples as myopathy. Those cases were difficult even for humans, as round small fibre atrophy was dominant in some ALS cases, as discussed above. This reflects the limitation in making a diagnosis based on morphology alone. Diagnostic accuracy would be improved by incorporating other pathological findings, such as necrotic fibres, internalized nuclei, regenerating fibres, and fibrosis, along with clinical findings and autoantibodies, into our model.

This single-center retrospective study, which primarily focused on the quantitative characterization of morphological features in relation to clinical diagnosis, has the following limitations. Clinical information was retrospectively reviewed and not standardised. In particular, we did not exclude the diagnosis of PM, which is not commonly used in current clinical practice. Two patients exhibiting both myopathy and neuropathy were excluded from the morphometric analysis. While we assume that mixed pathology would display a combination of the neuropathic and myopathic features identified in this study, further investigation in larger cohorts is needed to explore the potential synergistic effects of both aetiologies. We did not adjust for age or sex when defining small/large and/or round/angular fibres because these factors did not correlate with CSA or circularity in normal muscles (Table S1). However, a more precise morphological definition is expected to be established by analyzing a larger number of normal samples. In muscle morphometry, muscle fibres were filtered using an eccentricity threshold to exclude obliquely sectioned fibres. However, it remains a challenge for the software to completely exclude slightly obliquely sectioned fibres; therefore, investigators need to review the visualised images to confirm that fibres are properly segmented.

In conclusion, we quantitatively refined morphological and spatial features of biopsied muscles in myopathies and neuropathies using a machine learning-based software for muscle morphometry. We expect that this open-source tool will facilitate objective and efficient investigation of muscle histology.

## ACKNOWLEDGMENTS

We would like to thank Dennis W. Dickson (Mayo Clinic, Jacksonville) for his insights and meaningful comments and Yuka Nakajima (Institute of Science Tokyo) for histologic support.

## ETHICS STATEMENT

This retrospective study was approved by Institute of Science Tokyo Bioethics Committee (M2022-015) and was conducted based on comprehensive consent forms at the time of patients’ admission and an opt-out approach with disclosure of the study content on a hospital’s website and bulletin boards. Individual consent forms for this study were waived after the review by the ethics committee.

## AUTHORS’ CONTRIBUTIONS

DO conceived the study concept and design, acquired data, annotated datasets, developed software, analysed data, and drafted the manuscript, HK reviewed clinical records, acquired data, annotated datasets, and drafted the manuscript. HK and TY reviewed the manuscript and supervised the study. All authors approved the final manuscript.

## DATA AVAILABILITY STATEMENT

The software developed in this study is available at https://github.com/onnonuro/mmmetry.git with sample images and tutorials of Jupyter Notebook. The other data and codes used in the current study are available from the corresponding author upon reasonable request.

## List of abbreviations

ALS: amyotrophic lateral sclerosis
AP: average precision
ASS: antisynthetase syndrome
AUC: area under the curve
CSA: cross-sectional area
DM: dermatomyositis
H&E: hematoxylin and eosin
IBM: inclusion body myositis
IIM: idiopathic inflammatory myopathies
IMNM: immune-mediated necrotizing myopathy
LightGBM: light gradient boosting machine
PM: polymyositis
SD: standard deviation
WSI: whole slide image/imaging
YOLOv8: you only look once version 8

## Supplementary Material

**FIGURE S1.**
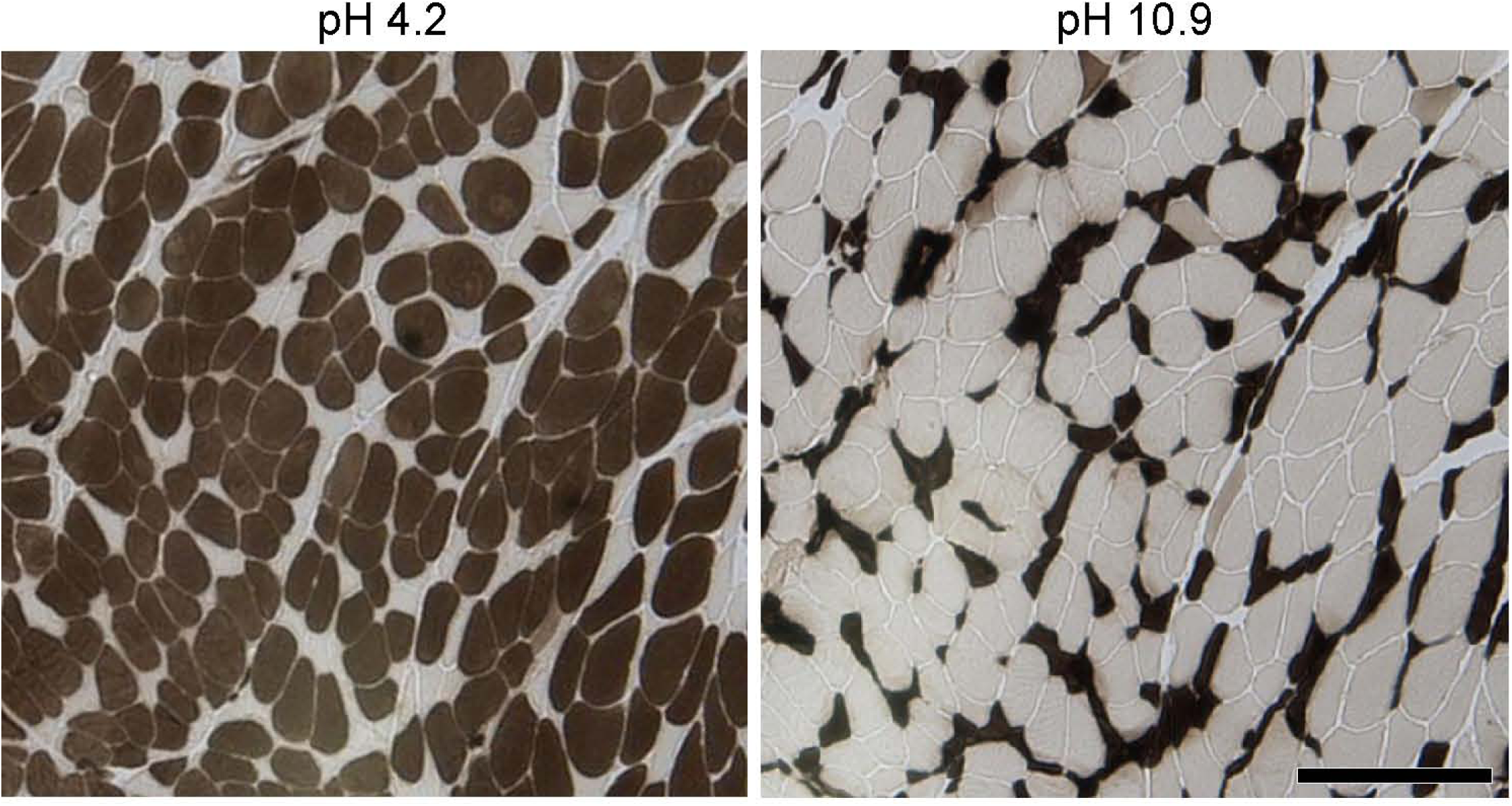
ATPase staining in a myopathy case with small angular fibers. ATPase staining images at pH 4.2 (left) and pH 10.9 (right) from the case of anti-MDA5 antibody positive dermatomyositis shown in Figure 3D. These images show that most of the small angular fibers are type 2, characterized by dark staining at pH 10.9. Scale bars: 1 mm.

**TABLE S1.** Correlation of morphometry with age and sex in normal samples.

|  | Sex |  |  | Correlation with Age |  |
| --- | --- | --- | --- | --- | --- |
|  | Male (n = 6) | Female (n = 14) | P value | Rho | P value |
| Mean CSA ($\mu\text{m}^2$) | 3078 [2488, 3601] | 3213 [2490, 4158] | 0.84 | 0.017 | 0.94 |
| Mean circularity ($\times 10^{-2}$) | 68.1 [67.9, 68.3] | 68.9 [67.8, 69.9] | 0.21 | -0.111 | 0.64 |

Values are presented as median [25th, 75th percentiles]. Statistical differences between sexes were assessed using the Mann-Whitney U test. Pearson correlation coefficients with age are presented as Rho. CSA, cross-sectional area.

**TABLE S2.** Muscle morphometry and spatial analysis in idiopathic inflammatory myopathies.

|  | Overall | IMNM | IBM | DM | ASS | PM | P value |
| --- | --- | --- | --- | --- | --- | --- | --- |
| *n* |  | 10 | 8 | 8 | 3 | 4 |  |
| Antibodies | | Anti-SRP(n=4),<br>Anti-HMGCR(n=3) | Anti-cN1A (n=3) | Anti-MDA5(n=3), Anti-TIF1-γ(n=2),<br>Anti-Mi-2(n=1),<br>Anti-NXP- 2(n=1) | Anti-Jo-1(n=3) | | |
| Onset to biopsy (month) | 5 [2, 33] | 7 [2, 14]<sup>IB</sup> | 71 [44, 95]<sup>DM,IM</sup> | 4 [2, 6]<sup>IB</sup> | 3 [2, 3] | 3 [2, 5] | **<0.01** |
| Number of fibres | 2931 [1101, 3833] | 3182 [1250, 3613] | 620 [372, 1377]<sup>DM</sup> | 3074 [2583, 3213]<sup>IB</sup> | 3833 [3024, 6234] | 4451 [4320, 4523] | **<0.01** |
| CSA ($\mu\text{m}^2$) | | | | | | | |
| Mean | 2693 [1888, 3866] | 2488 [2051, 2768]<sup>IB</sup> | 4931 [4367, 5850]<sup>IM</sup> | 2872 [2183, 3577] | 1888 [1673, 2053] | 1738 [1233, 2445] | **<0.01** |
| SD | 1230 [877, 2057] | 1188 [945, 1445]<sup>IB</sup> | 3530 [2941, 4697]<sup>DM,IM</sup> | 1094 [798, 1448]<sup>IB</sup> | 877 [763, 1054] | 997 [673, 1334] | **<0.01** |
| Circularity ($\times 10^{-2}$) | | | | | | | |
| Mean | 70.9 [68.1, 73.0] | 71.2 [68.5, 72.5] | 68.4 [67.2, 73.8] | 69.9 [68.3, 71.7] | 71.0 [68.5, 72.2] | 72.1 [71.1, 73.0] | 0.78 |
| SD | 9.9 [8.6, 11.1] | 9.9 [8.3, 10.7] | 11.2 [10.4, 11.9] | 9.8 [9.2, 10.0] | 10.1 [9.2, 10.8] | 9.4 [8.9, 9.6] | 0.11 |
| Mean in small fibres | 68.8 [65.8, 70.7] | 69.0 [67.0, 71.1] | 68.7 [64.0, 73.2] | 66.7 [65.1, 67.4] | 68.8 [66.8, 69.7] | 70.0 [69.3, 71.1] | 0.43 |
| Mean in large fibres | 68.8 [65.7, 73.3] | 65.7 [62.1, 73.4] | 69.8 [68.7, 74.0] | 67.6 [66.1, 69.9] | 64.3 [61.3, 67.3] | 70.5 [63.8, 74.8] | 0.6 |
| Fibre population (%) |  |  |  |  |  |  |  |
| Small | 23.6 [11.5, 45.5] | 28.1 [17.2, 37.1] | 20.6 [13.4, 25.4] | 10.2 [7.6, 23.0] | 52.1 [35.7, 52.5] | 52.0 [30.3, 70.9] | 0.15 |
| Large | 4.5 [0.5, 30.0] | 3.4 [0.9, 9.6]<sup>IB</sup> | 41.8 [35.7, 48.4]<sup>DM,IM</sup> | 5.9 [0.5, 16.4]<sup>IB</sup> | 0.7 [0.3, 0.9] | 2.3 [0.1, 6.3] | **<0.01** |
| Round | 20.8 [15.0, 28.5] | 21.5 [17.7, 26.4] | 21.2 [15.2, 41.2] | 15.1 [13.6, 22.8] | 24.3 [18.4, 26.1] | 26.6 [22.8, 30.0] | 0.63 |
| Angular | 12.5 [8.5, 18.4] | 12.5 [6.9, 17.5] | 18.2 [12.3, 24.0] | 12.3 [8.9, 17.0] | 12.5 [9.3, 18.9] | 9.5 [7.7, 11.6] | 0.47 |
| Small angular | 3.41 [2.36, 5.58] | 3.42 [3.06, 5.01] | 3.83 [2.11, 5.52] | 2.40 [1.80, 3.92] | 8.71 [5.54, 12.17] | 5.41 [4.62, 6.99] | 0.45 |
| Small round | 4.48 [1.86, 9.00] | 5.34 [2.61, 8.30] | 5.14 [1.79, 12.10] | 1.62 [1.06, 2.07] | 5.69 [4.99, 7.35] | 9.15 [6.04, 14.50] | 0.23 |
| Large angular | 0.55 [0.09, 3.02] | 0.92 [0.11, 1.51] | 5.80 [4.28, 8.86] | 0.79 [0.12, 1.79] | 0.09 [0.05, 0.29] | 0.16 [0.06, 0.29] | **0.03** |
| Large round | 1.28 [0.08, 5.36] | 0.41 [0.07, 1.17]<sup>IB</sup> | 7.45 [5.95, 12.05]<sup>DM,IM</sup> | 0.74 [0.11, 2.16]<sup>IB</sup> | 0.00 [0.00, 0.09] | 0.77 [0.02, 2.32] | **<0.01** |
| Spatial analysis |  |  |  |  |  |  |  |
| Grouped fibres (%) | 32 [16, 63] | 33 [28, 47] | 22 [12, 32] | 26 [14, 57] | 77 [63, 82] | 84 [63, 93] | 0.07 |
| Nodes per cluster | 4.6 [3.9, 6.2] | 4.8 [4.1, 5.3] | 3.5 [3.2, 3.9] | 4.5 [3.8, 5.5] | 7.1 [6.6, 11.1] | 20.7 [13.2, 26.4] | **<0.01** |
| Edges per node | 2.0 [1.8, 2.5] | 2.1 [2.0, 2.2] | 1.7 [1.5, 1.9] | 1.9 [1.8, 2.2] | 2.5 [2.4, 3.0] | 3.9 [3.2, 4.3] | **0.01** |

Values are presented as median [25th and 75th percentiles]. Statistical differences were evaluated using the Kruskal–Wallis test and presented as P values. The pairwise Steel–Dwass test was followed for variables observed with significant differences. Superscripts of IBM, DM, and IMNM mean $P < 0.05$ vs. IBM, DM, and IMNM, respectively. ARS, aminoacyl transfer RNA synthetase; ASS, antisynthetase syndrome; CSA, cross-sectional area; cN1A, cytosolic 5′-nucleotidase 1A; DM, dermatomyositis; HMGCR, 3-hydroxy-3-methylglutaryl-coenzyme A reductase; IBM, inclusion body myositis; IMNM, immune-mediated necrotizing myopathy; MDA5, melanoma differentiation-associated gene 5; NXP2, nuclear matrix protein 2; PM, polymyositis; SD, standard deviation; SRP, signal recognition particle; TIF1γ , transcriptional intermediary factor 1γ .

